# The Index of Number of Events and Severity (INES): A Novel Instrument for Quantifying Lifetime Illness Burden in Bipolar Disorder

**DOI:** 10.64898/2026.08.06.26358032

**Authors:** Inés Carpio-López, Inés García-Ortiz, Diego Romero-Miguel, Elisabet Madridejos- Palomares, Laura Jiménez-Muñoz, María P. Rodríguez-Gómez, Lucía Albarracín-García, Enrique Baca-García, Claudio Toma

## Abstract

Bipolar disorder (BD) is a chronic psychiatric condition affecting approximately 1–2% of the population, characterized by depressive and manic episodes. BD comprises two main subtypes, defined by the presence of mania (BD-I) or hypomania (BD-II). Commonly used clinical scales, including the Global Assessment of Functioning (GAF), Clinical Global Impressions (CGI), and World Health Organization Disability Assessment Schedule (WHODAS), assess functional impairment at the time of evaluation. However, they may not adequately capture cumulative lifetime illness burden or provide a retrospective measure of clinical severity.

Here, we introduce the Index of Number of Events and Severity (INES), a novel instrument designed to quantify longitudinal illness-course severity in BD by integrating cumulative clinical events with illness duration.

INES incorporates psychosis and rapid cycling as dichotomous variables and quantifies hospitalizations, suicide attempts, and affective episodes as discrete categories. INES was evaluated in 307 individuals from the MadManic cohort. It correlated moderately with GAF and CGI, while its strongest association was observed with WHODAS (r=0.347). Factor analysis over the four scales supported a two-factor structure, where INES loaded alongside WHODAS, capturing the variability of structured instruments. Linear modelling indicated that traditional scales explained only 14.4% of the variance of INES, suggesting that this scale captures clinical information largely unaccounted by the other instruments. INES was the only to differentiate between BD subtypes, with higher severity observed in individuals with BD-I. These findings support INES as a reproducible tool for capturing cumulative lifetime severity in BD, with potential utility in clinical and genetic studies.

## 1. INTRODUCTION

Bipolar disorder (BD) is a chronic psychiatric condition affecting approximately 1–2% of the population. It is characterised by mood shifts between depressive and manic episodes (Nierenberg et al., 2023). BD comprises two main subtypes based on the manic expression: BD-I, marked by full manic episodes and associated with greater clinical severity; and BD-II, defined by hypomanic episodes (Tondo et al., 2022).

Although BD-I and BD-II share substantial genetic liability (r=0.829), recent evidence indicates that BD-I shows higher genetic correlation with schizophrenia, whereas BD-II overlaps mostly with major depressive disorder, panic disorder, and other internalizing traits (O’Connell et al., 2025; Veen et al., 2026).

In clinical practice, several scales are commonly used to capture functional impairment or severity in individuals with BD. These scales include: i) the Global Assessment of Functioning (GAF) (Endicott, 1976), a clinician-rated scale ranging from 0 to 100 that provides an overall assessment of functioning; ii) the Clinical Global Impressions (CGI) scale (Spearing et al., 1997), which similarly provides a clinician rated 1–7 impairment score; and iii) the World Health Organization Disability Assessment Schedule (WHODAS) (Guilera et al., 2015; Üstün et al., 2010), which provides an objective multidomain assessment of functioning and disability over the previous 30 days.

These instruments typically capture disease state at the time of the assessment rather than quantifying overall lifetime illness burden. When none of these scales is available, especially in genetic studies, age of onset (AOO) is usually used as a proxy of severity (Toma et al., 2018), although it is not clear how much variability the AOO is able to capture from the whole functional disability or severity.

Thus, a valid instrument to retrospectively measure severity burden could be useful at clinical level for prognostic assessment of functional impairment (Kapczinski et al., 2008), and at genetic level for association with polygenic risk scoring and highly disruptive genetic variants (Toma, 2020; Toma et al., 2018).

Dissecting heterogeneous phenotypes such as BD in distinct clinical features or traits may increase power to identify genetic risk profiles. Indeed, a recent study shows that AOO, BD subtype, psychotic symptoms, and manic severity are clinically relevant dimensions that may capture genetic heterogeneity in BD (Van Loo et al., 2023). Thus, the administration of a standardised instrument that capture retrospective illness burden could therefore facilitate more precise genotype-phenotype analysis and improve characterization in genetic and clinical studies.

Here, we present the Index of Number of Events and Severity (INES), an instrument designed to quantify illness-course severity in BD by integrating the cumulative burden of clinical events with illness duration.

## 2. METHODS

### 2.1. Cohort, clinical data, and variables for the INES

The INES was validated using data from 307 BD cases from the Madrid Manic cohort (MadManic), a Spanish BD cohort with extensive clinical data, including socio-demographic data, biometric measures, detailed illness trajectories, comorbidities, hospitalizations and suicidality, enabling a comprehensive lifetime clinical assessment (García-Ortiz et al., 2026).

INES combines two dichotomous and six polytomous variables. The dichotomous variables, which include psychosis and rapid cycling, were coded as 0 when absent and 3 when present. The polytomous variables comprised number of hospitalizations, number of suicide attempts, and number of manic, hypomanic, depressive, and mixed episodes. Each variable was coded on a four-level ordinal scale, with 0 indicating no events, 1 indicating one to two events, 2 indicating three to five events, and 3 indicating more than five.

To balance the influence of disease duration and the cumulative number of clinical events, INES combines two equally weighted components: one adjusted for disease duration and one unadjusted. This approach prevents excessive penalization of longer illness duration. A logarithmic transformation, followed by rescaling to a 0–10 range, was applied to reduce skewness and facilitate interpretation, with higher scores indicating greater lifetime illness severity. The INES script for score calculation is available at https://github.com/madpsych/INES.git.

INES scores were calculated in the cohort using *R* software (v4.4.1) (R Core Team, 2024). **Table 1** reports the number of individuals with BD included in the study, stratified by subtype, for whom INES, GAF, CGI, and WHODAS 2.0 (32 items) data were available.

**Table 1.**
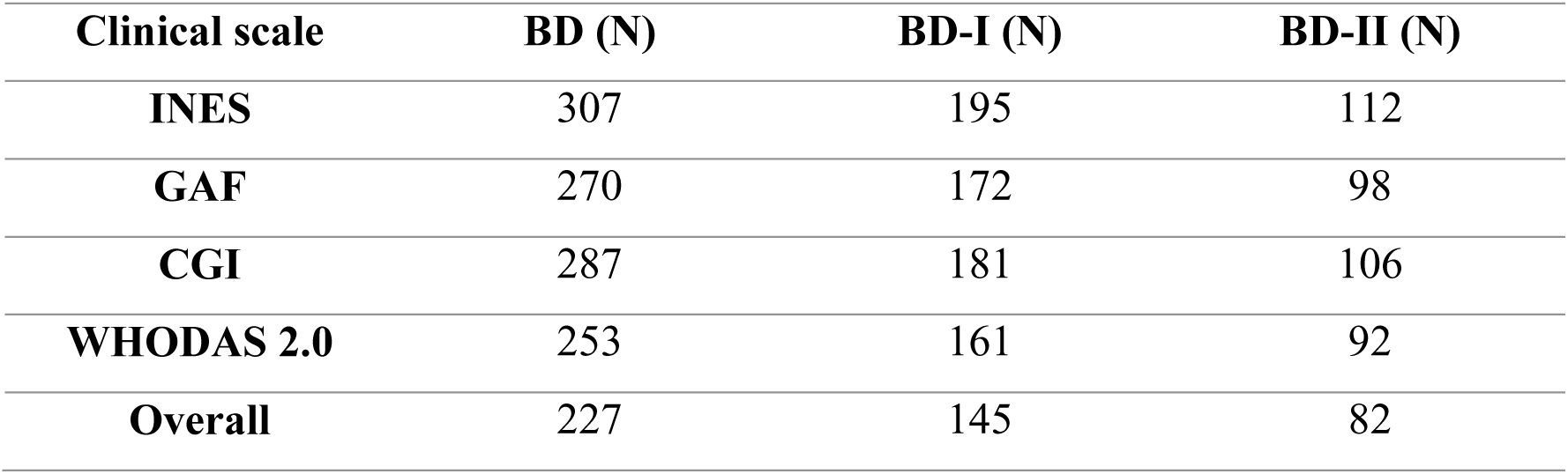
Number of BD subjects from the MadManic cohort with available scores of INES, GAF, CGI, WHODAS 2.0 (32 items). Additional information is also provided for BD-I and BD-II subtypes.

| **Clinical scale** | **BD (N)** | **BD-I (N)** | **BD-II (N)** |
| --- | --- | --- | --- |
| **INES** | 307 | 195 | 112 |
| **GAF** | 270 | 172 | 98 |
| **CGI** | 287 | 181 | 106 |
| **WHODAS 2.0** | 253 | 161 | 92 |
| **Overall** | 227 | 145 | 82 |

### 2.2. Statistical analyses

Pairwise correlations among INES, GAF, CGI and WHODAS were computed to assess whether INES aligns with existing measures of clinical severity and functioning. An exploratory factor analysis (EFA) was performed using *psych* package (v2.6.3) (Revelle, 2026) to examine the latent structure underlying INES and the three established clinical instruments and identify shared or distinct dimensions of severity and functioning (Watkins, 2018). Prior to factor extraction, the Kaiser-Meyer-Olkin (KMO) measure and Bartlett’s sphericity test were computed to evaluate the adequacy of the correlation matrix for factor analysis (Bartlett, 1950; Kaiser, 1970). A parallel analysis supported a two-factor solution, which was then fitted using: i) maximum likelihood extraction to calculate the factorial loadings of each scale, reflecting their contribution to the latent factor; ii) and *oblimin* rotation, allowing the latent factors to be correlated. To further assess model adequacy, a one-factor EFA model including the four scales was also fitted for comparison with the two-factor model. Communality was calculated to address the extent to which each scale shared variance with the other instruments.

Additionally, to quantify how much of the variance of INES scale was explained by the other clinical instruments, we fitted a linear regression model with INES as the outcome and WHODAS, GAF, and CGI as predictors. Relative importance analyses were performed using the *relaimpo* package (v2.2.7) (Grömping, 2006) to estimate the individual contribution of each instrument to the total variance of INES accounted by the model.

To determine whether any of the clinical measures could distinguish between BD-I and BD-II subtypes, the four scales were compared using a Wilcoxon rank-sum test with 10,000 permutations (Hothorn et al., 2008). To evaluate whether INES provided incremental information for predicting BD subtype beyond the existing clinical measures, we then fitted and compared two logistic regression models with BD subtype as the outcome: a reduced model including GAF, CGI, and WHODAS, and a full model additionally including INES.

## 3. RESULTS

### 3.1. INES calculation and clinical data inclusion

The INES score was calculated as follows:

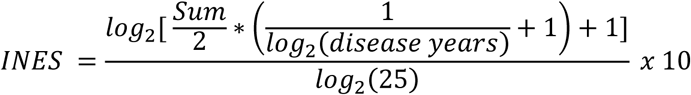

The term *Sum* refers to the total score obtained by adding: i) lifetime presence of both psychosis and rapid cycling clinical variables, coded as binary items (yes=3; no=0; maximum score=6); ii) number of hospitalizations, suicide attempts, and manic, hypomanic, depressive, and mixed episodes, on a four-level ordinal frequency scale from 0 to 3 (none=0; 1–2 times=1; 3–5 times=2; >5 times=3; maximum score=18). Thus, *Sum* ranged from 0 to 24. *Disease years* was defined as the number of years elapsed from the first mood episode to the time of last assessment. Because INES is designed to capture longitudinal severity rather than early-stage clinical presentation, we applied the index only to individuals with at least one year of illness duration, as criteria to be used in future applications of the scale. The calculation of INES was performed in 307 BD patients, where their scores ranged from 2.47 to 8.09 (**Fig. 1A**).

**Figure 1.**
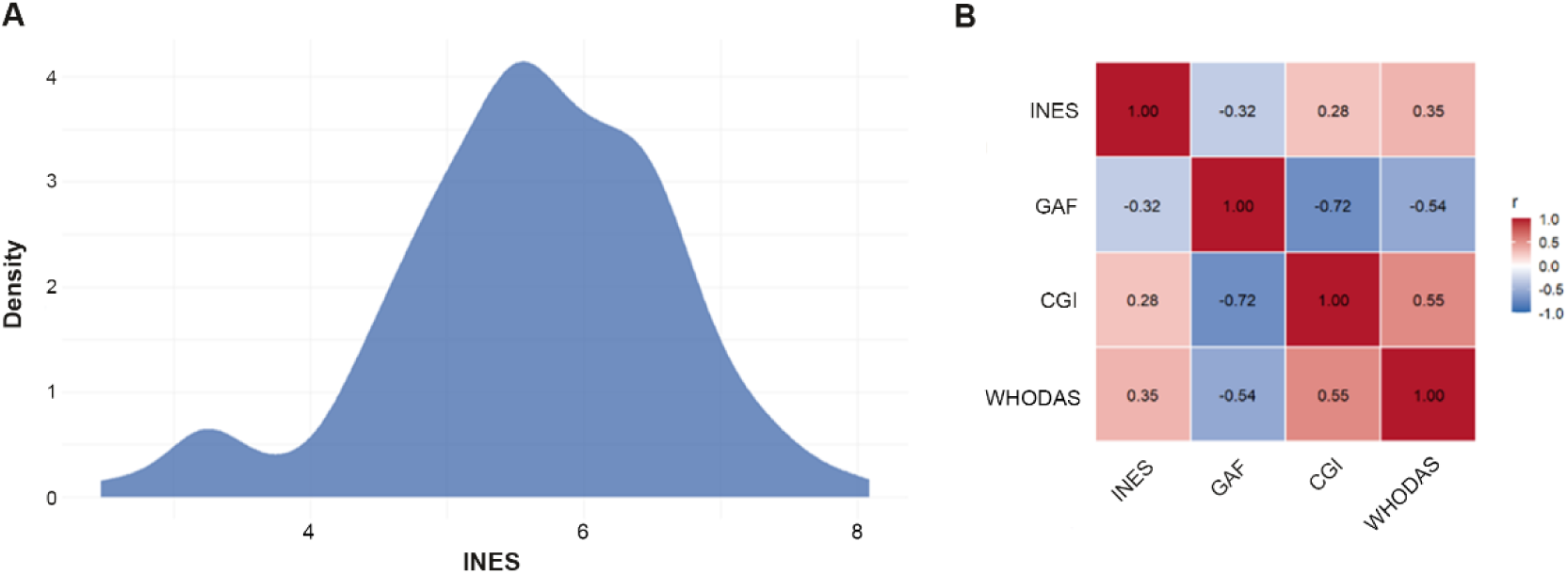
INES distribution and correlations with additional clinical scales capturing functional disability or severity. A) Density distribution of INES scores in the MadManic cohort (N=307 BD individuals). B) Pearson correlation heatmap between INES, GAF, CGI, and WHODAS. Red indicates positive correlations and blue indicates negative correlations.

### 3.2. Comparison of INES with other clinical scales

Pairwise correlations across the four scales showed that INES presented moderate correlations with the other clinical instruments, being slightly higher with WHODAS (r=0.347; *p*=1.42E-08) than GAF (r=-0.316; *p*=1.09E-07) or CGI (r=0.279; *p*=1.55E-06). GAF and CGI were strongly correlated with each other (r=-0.719; *p*=3.36E-44), as expected given their conceptual similarity and reliance on clinical assessment. Correlations involving GAF were negative, as GAF scores are inverted, with higher scores indicating lower functional disability (**Fig. 1B**).

The adequacy of the correlation matrix for factor analysis was supported by the KMO statistic, which indicated an acceptable measure of sampling adequacy (MSA = 0.73), and by Bartlett’s test of sphericity, which was highly significant (*p*=1.81E-61). Consistent with the observed correlations, the two-factor EFA revealed a coherent and interpretable latent structure. The first maximum-likelihood factor (ML1), defined by GAF and CGI, reflected scales based on subjective clinician evaluation, whereas the second factor (ML2), containing WHODAS and INES, implicated scales based on structured instruments of objective measures. CGI and GAF showed strong factor loadings on ML1 (λ=0.90 and λ=-0.78, respectively), while WHODAS and INES showed moderate factor loadings on ML2 (λ=0.46 and λ=0.59, respectively) (**Fig. 2A**). The two factors were positively correlated (r=0.69). Together, they accounted for 56.8% of the total variance, with ML1 and ML2 accounting for 40.3% and 16.5%, respectively. Communalities were high for GAF (h²=0.717) and CGI (h²=0.753), moderate for WHODAS (h²=0.494), and lowest for INES (h²=0.307), suggesting that INES successfully captures information on clinical course that is only partly shared with the other scales.

**Figure 2.**
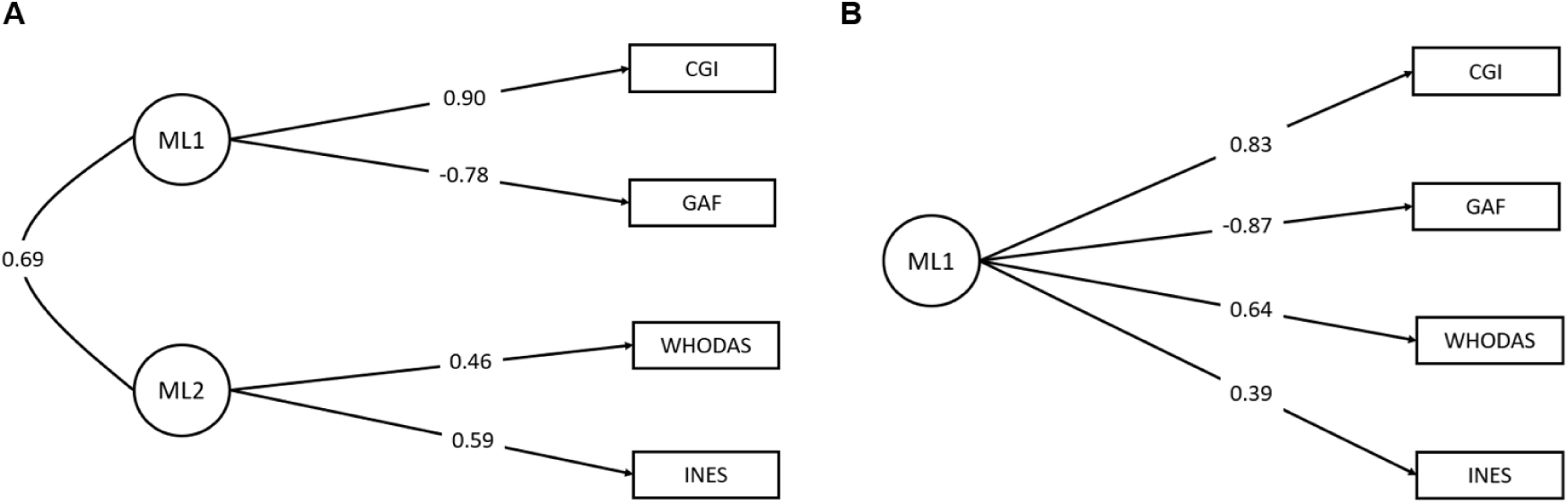
Exploratory factor analysis (EFA) diagrams. Numbers alongside the arrows indicate the standardized factor loadings for each clinical scale to their assigned factor. A) Two-factor model; the correlation between the latent factors (ML1 and ML2) is also shown. B) One-factor model.

A one-factor EFA model was also calculated to determine whether the four instruments could instead be represented by a single general factor. In this analysis, GAF and CGI showed the strongest factor loadings (λ=-0.87 and λ=0.83, respectively), whereas WHODAS had a moderate loading (λ=0.64) and INES showed the weakest loading (λ=0.39) (**Fig. 2B**). In this case, the one-factor model accounted for 50.2% of the variance, which was lower than the variance explained by the two-factor solution.

The limited overlap between INES and current clinical measures was further supported by linear modelling, in which WHODAS, GAF, and CGI jointly accounted for only 14.4% of the variance in INES (R²=0.144). WHODAS was the only scale significantly associated with INES (*p*=9.68E-04). In line with these results, relative importance analysis showed that WHODAS was the main contributor, accounting for 48.5% of the explained variance, followed by GAF with 30.5% and CGI with 21%. Thus, 85.6% of INES variance remained unexplained, indicating that most of the information captured by INES is not reflected in current clinical instruments.

Finally, INES was the only instrument that significantly distinguished between BD-I and BD-II types in permutated Wilcoxon rank-sum test (*p*=0.027), showing higher scores in BD-I, whereas the other clinical scales were unable to discriminate between BD subtypes: GAF (*p*=0.36), CGI (*p*=0.76) and WHODAS (*p*=0.92). To determine whether the observed INES difference provided incremental information for BD subtype prediction beyond current clinical instruments, reduced and full logistic regression models were subsequently compared. The reduced logistic regression model, which included GAF, CGI, and WHODAS, had negligible explanatory power for BD subtype (pseudo-R^2^=0.0017), and none of its predictors was statistically significant (*p*=0.49-0.80). In the full model, the inclusion of INES increased the explanatory power (pseudo-R²=0.0071) with an increase of the explained variance of 0.0054. Although the INES estimate in the logistic model pointed in the expected direction (β=1.917), indicating higher scores in BD-I, it did not reach statistical significance (*p*=0.21). These findings suggest that this instrument was able to discriminate between BD-I and BD-II at the group level; however, it was not sufficient for accurate subtype prediction at the individual level.

## 4. DISCUSSION

INES was specifically designed to address a major limitation in psychiatric research, accounting for the lack of standardized instruments capable of retrospectively quantifying cumulative illness burden using routinely collected clinical data. Current clinical scales such as GAF, CGI, and WHODAS primarily capture symptom severity at the time of assessment, providing limited insight into long-term disease trajectory (Endicott, 1976; Guilera et al., 2015; Spearing et al., 1997; Üstün et al., 2010). We present INES, a novel instrument that aims to fill this gap by quantifying lifetime severity through the integration of the number of major clinical events together with illness duration. This approach aligns with allostatic load models of BD, where repeated affective episodes and stressors progressively contribute to long-term burden and functional decline (Kapczinski et al., 2008).

INES captures a clinical dimension related to existing scales, as showed by correlations with GAF, CGI, and WHODAS, but also presents substantial differences. The two-factor EFA model, derived by the parallel analysis, explained more variance than the one-factor model, providing a better representation of the relationship amongst scales. Under this model, one of the factors clustered INES and WHODAS together, supporting that these scales capture part of the same information, while the other factor grouped GAF and CGI, which strongly depend on subjective clinical evaluation. Moreover, INES showed the lowest communality amongst the four instruments, suggesting that it successfully captures information on clinical course that is only partly shared by the other scales. This interpretation is consistent with the limited variance of INES explained by CGI, GAF, and WHODAS, indicating that this novel instrument is not redundant compared with existing scales, but able to capture an independent dimension of clinical severity.

Additionally, INES was also the only instrument that was able to discriminate between BD subtypes. BD-I cases displayed higher INES scores than BD-II, consistent with the more disruptive long-term course typically associated with full manic episodes (Tondo et al., 2022). On the other hand, INES was not able to predict BD subtype at individual-level, which is expected considering that additional variables and instruments would be needed in order to disentangle heterogeneous phenotypes such as BD. However, the design of this scale focuses on evaluating long-term severity burden, rather than serving as a diagnostic instrument.

Importantly, the quantification of retrospective illness performed by INES may be particularly valuable in genetic studies, where standardized severity measures are often scarce or rely on scales focused on specific temporal assessments, which is not ideal for quantitative genetic analyses (Van Loo et al., 2023).

A limitation of INES is its strong reliance on retrospective clinical records, which are not always available.

In conclusion, INES is a novel quantitative instrument that captures lifetime illness severity in BD by integrating cumulative clinical events with disease duration. Unlike current available scales focused on functioning or disability at time of the assessment, INES provides a structured and retrospective measure of long-term burden. Our findings show that INES represents a largely independent dimension of severity and offers a practical tool for genetic and clinical research.

## Data Availability

All data produced in the present study are not publicly available. However, requests can be sent to the authors for individual assessment.

## CRediT AUTHORSHIP CONTRIBUTION STATEMENT

**Inés Carpio-López:** Methodology, Investigation, Formal Analysis, Data Curation, Conceptualization, Writing – review – editing, Writing – original draft. **Inés García-Ortiz:** Methodology, Investigation, Formal Analysis, Data Curation, Conceptualization, Writing – review – editing, Writing – original draft. **Diego Romero-Miguel:** Methodology, Conceptualization, Writing – review – editing. **Elisabet Madridejos-Palomares:** Data Curation, Visualization, Validation. **Laura Jiménez-Muñoz:** Data Curation, Visualization, Validation. **María P. Rodríguez-Gómez:** Data Curation, Visualization, Validation. **Lucía Albarracín-García:** Data Curation, Visualization, Validation. **Enrique Baca-García:** Visualization, Validation, Supervision, Resources, Funding. **Claudio Toma:** Data Curation, Supervision, Resources, Funding, Methodology, Conceptualization, Writing – review – editing.

## DECLARATION OF COMPETING INTEREST

EBG has been a consultant for or has received honoraria or grants from Janssen Cilag, Lundbeck, Otsuka, Pziffer, Servier, Deprexis and Sanoffi. No other disclosures are reported from the other authors.

## ACKNOWLEDGMENTS

We thank Daniel García Rubio for his mathematical support during the development of the INES. This study was supported by grants: RyC2018-024106-I, PID2020-114996RB-I00, CNS2022-135318, PID2023-149154OB-I00 funded by MICIU/AEI/10.13039/501100011033, FEDER UE, European Union NextGenerationEU/PRTR and ESF Investing in your future (to Toma). Additional support for this study was received by CIBER-Consorcio Centro de Investigación Biomédica en Red (CB/07/09/0025), the Instituto de Salud Carlos III with the support of the European Regional Development Fund (ISCIII PI23/00614; PMP24/00026), Fundació La Marató de TV3 (202226-31) and by CaixaResearch Health 2023 LCF/PR/HR23/52430033 (to Baca-Garcia).

The Centro de Biología Molecular Severo Ochoa (CBM) is supported by the Fundación Ramón Areces and holds the Severo Ochoa Centre of Excellence distinction (MICIN, CEX2021-001154-S). García-Ortiz was supported by the Fundación Tatiana Pérez Guzmán el Bueno fellowship, and Dr Romero-Miguel by the Juan de la Cierva fellowship (grant JDC2023-052237-I) funded by MCIN/AEI/10.13039/501100011033 and ESF+. The funders had no role in the design collection, management, analysis, and interpretation of the data.

## Notes

### Clinical Protocols

https://github.com/madpsych/INES.git

### Author Declarations

Approvals to handle clinical or behavioural scales of BD patients and controls were obtained by the research ethics committee of the Fundacion Jimenez Diaz Hospital (15/21; 11/23) and the Spanish National Research Council (Consejo Superior de Investigaciones Cientificas, CSIC) (13/2021; 109/2023; 030/2025). All participants provided written informed consent.

